# Health insurance in Nigeria: Findings from the People’s Voice Survey

**DOI:** 10.64898/2026.03.03.26347555

**Authors:** Kevin Croke, Chike Nwangwu, Olufunke Fasawe, Ifeyinwa Aniebo, Karima Ladhani, Margaret E. Kruk, Oyebanji Filani

## Abstract

The recent Lancet Commission on Nigeria’s health system highlighted high out of pocket expenditures on health and underfunding of the public health sector as major obstacles to Nigeria’s achievement of the Sustainable Development Goals. Nigeria has sought to address these gaps by extending health insurance coverage. This paper measures health insurance coverage and access to care in Nigeria circa 2023, using the first round of the People’s Voice Survey (PVS). We analyze health insurance coverage by calculating coverage rates and using multivariate logistic regression to estimate associations between insurance coverage, socioeconomic characteristics, and health system utilization. In 2023, only 2% of Nigerians had insurance from the National Health Insurance Scheme; higher education and higher income levels were the most notable predictors of NHIS access. Chronic illness and self-reported health were not associated with insurance status. Respondents with insurance were less likely to use public sector primary care providers as their usual source of care, and were more likely to use private hospitals. Those with insurance are also more likely to have had an inpatient hospitalization in the preceding year, and more likely to have received key preventive screenings. While those with insurance receive more and better care in Nigeria, insurance access has been limited to relatively advantaged population groups. Rapid mobile phone-based surveys such as PVS could help policymakers in Nigeria track insurance coverage and whether it contributes to reversal of these trends over time.

## Introduction

Nigeria is a critical country for attainment of the Sustainable Development Goals (SDGs), including SDG target 3.7 (universal health coverage). With over 215 million inhabitants, Nigeria has the largest population in sub-Saharan Africa, and accounts for significant fractions of global under 5 and maternal mortality.^1^

A recent Lancet Commission highlighted several key challenges facing Nigeria’s health system, including underfunding of the health sector (the percentage of GDP spent on health is among the lowest in the world),^2^ and high out of pocket expenditures (OOPE) for those seeking care. The Commission called for a scale up of health insurance, with public financing of insurance coverage for poor Nigerians.^1^ However, as of 2018, the Nigeria Demographic and Health Survey showed that only 3% of adults aged 15-49 had any form of insurance coverage.

To address these gaps, Nigerian policymakers have initiated major health reforms. The 2014 National Health Act has been more recently bolstered by the National Health Insurance Authority Act (2022), both of which have had health insurance as their focus.^3^ Nigeria is also currently undergoing additional reforms including the National Health Sector Renewal Investment Initiative (NHSRII), led by the current coordinating Minister of Health and Social Welfare Dr. Muhammad Pate; these reforms also support Nigeria’s efforts towards UHC.

Globally, Nigeria’s experience is highly relevant. A large number of LMICs are currently attempting to scale up health insurance programs to achieve universal health coverage. Yet there are important global policy debates about the conditions under which these approaches are likely to succeed.^4^ While contributory insurance programs can complement tax-financed systems, they can also lead to a range of issues including fragmentation, inequity, and increased costs due to provider-initiated demand.^5^ Careful evaluation of the impact and distributional implications of Nigeria’s expanding health insurance programs is therefore warranted, and likely to be informative to other countries in the region and globally. This study uses 2023 People’s Voice Survey data to identify the levels and determinants of health insurance coverage in Nigeria, and the implications of this coverage for health service access and utilization.

### Background: National health insurance in Nigeria

Since independence, Nigeria’s health system was based on public financing and provision of care through hospitals and health centers run by the federal government, states, and local government authorities. However over time, gaps in public provision led to the emergence of a growing private sector, and accordingly, increased out of pocket expenditures on health services. The National Health Insurance Scheme (NHIS) then emerged as a response to this growing burden of out-of-pocket expenditure.

The NHIS Act was first signed into law in 1999 by President Olusegun Obasanjo. The scheme, launched in 2005, had the goal of providing health insurance coverage (UHC) for all Nigerians by 2015 *(“universal coverage and access to adequate and affordable healthcare in order to improve the health status of Nigerians, especially for those participating in the various programs of the Scheme”)*. It also aimed to formally integrate the private sector into health service delivery by including private healthcare facilities. The scheme was launched as a collaborative project between the NHIS, health management organizations (HMOs), and healthcare providers. The NHIS accredits HMOs as purchasers, and healthcare facilities as service providers, to provide the defined benefits package of services to enrolled Nigerians. HMOs then purchase services for enrolled beneficiaries on behalf of the NHIS. The NHIS envisioned three main programs covering the formal and informal sectors: the Formal Sector Social Health Insurance Program (FSSHIP) for employees in the formal sector (public and private), funded through percentage employee-based contributions; the Urban Health Self-employed Social Health Insurance Program (USSHIP); and the Rural Community Social Health Insurance Program (RCSHIP). The latter two were for the informal sector and funded by non-voluntary flat-rate premiums based on selected health benefits package chosen by the enrollee. Contributions for the FSSHIP were based on employer-employee contributions distributed as 10% of basic salary from the employer, and 5% of basic salary from the employee.^6^

Despite the ambitious plan to reach Universal Health Coverage by 2015, several studies have reported low coverage of the NHIS.^7,8^ Although multiple reasons could be adduced, three key challenges stood out: (a) The NHIS did not have a mandatory element, which limited its ability to scale, (b) The program had limited ability to aggregate and pool funds from large sections of the population that worked and operated in informal sectors, (c) challenges to governance in the NHIS, with a high turnover of chief executives (6 in 10 years).^6^ Previous studies have reported that most of those covered under the scheme are federal government employees and their dependents (via the Formal Sector Social Health Insurance Program, which was established to cover employees of federal, state, and local governments and those of private institutions employing at least ten workers).^9^ Even FSSHIP faced multiple challenges in implementation.^9^ Additional challenges for the NHIS included unwillingness by state government employees to make employee contributions to the scheme, concerns by healthcare providers that the amount of the capitation paid was insufficient, inadequate funding due to the low adoption, poor quality of drugs and services, limited accountability and accusations of corruption, and extremely low coverage of the informal sector.^10,11^

A major reform which focused in part on supporting health insurance and universal health coverage took place in 2014, when Nigeria’s National Health Act (NHA) was passed. A key component of this reform was the creation of the Basic Health Care Provision Fund (BHCPF), which aimed to allocate 1% of Nigeria’s consolidated revenue to this new health fund.^12^ The BHCPF, which was first launched in 2019, then redesigned in 2023 as the BHCPF 2.0, operates through three “gateways”: the NHIS, the National Emergency Medical Treatment, and the National Primary Health Care Development Agency (NPHCDA). All three gateways aim to provide a basic minimum package of health services (BMPHS) that beneficiaries can access, with the NHIA specifically providing health finance security to enable equitable health service delivery and mitigate catastrophic out-of-pocket expenditure^3^.

Due to the chronic systemic challenges that the NHIS faced and the inability to achieve universal health coverage after over a decade, there were calls for reform of the NHIS, leading to the signing of the new National Health Insurance Authority (NHIA) Act.^13^ On May 19, 2022, this new legislation repealed the NHIS and enacted the National Health Insurance Authority. The NHIA’s revised mandate includes regulating state-level health insurance schemes, ensuring that health insurance is mandatory for all Nigerians, and enforcing the basic minimum package of health services for Nigerians across federal, state, and private health insurance schemes.^13^ In addition to creating the NHIA, the Act also permits the establishment of state health insurance schemes and establishes the Vulnerable Groups Fund to provide fully subsidized healthcare service coverage for vulnerable persons through the Basic Healthcare Provision Fund.

With the reform of the NHIS, it has been expected that Nigeria could accelerate universal health coverage for all Nigerians. However, ongoing monitoring and analysis to track progress is needed. This paper uses a new data source (the People’s Voice Survey) to shed light on key elements of health system performance, including questions of health financing, insurance coverage, and access to care. The PVS has been used to study health system performance across multiple domains including primary health care (PHC), insurance coverage, health system confidence, and vaccination across multiple countries.^14,15,16^ Single country analyses using PVS data have focused on user perceptions of quality and public versus private sector use in Mexico,^17^ and utilization patterns and coverage of key health system preventive services in Laos.^18^ One notable use of PVS has been to identify challenges facing health insurance programs across low- and middle-income countries (LMICs), as previous research has demonstrated that across five LMICs, public health insurance is only weakly associated with access to health services.^19^

In this paper we focus on the PVS measurement of health insurance status and health utilization. In Nigeria, recent major health reforms including National Health Act (2014) and National Health Insurance Authority Act (2022) have health insurance coverage at their center, yet have not been fully evaluated. Previous research on Nigeria’s health system has highlighted very high OOPE for health, as well as low public sector spending. The PVS has measured insurance status and access to services, and described heterogeneity by geographical and individual characteristics. As such, data on health insurance coverage and access to services from the PVS can help to establish a baseline against which current efforts to increase coverage can be evaluated.

## Materials and Methods

The PVS is a new primarily mobile phone-based survey designed to integrate people’s voices into health system measurement. It has been conducted using a random digit dialing sampling frame in 15 countries to date.^20^ The PVS in Nigeria was implemented in June and July 2023, with total sample size of 2,555. Interviews were conducted in Hausa, Igbo, Pidgin, Yoruba, and English. While mobile phone surveys typically oversample higher socioeconomic status groups, the PVS data was reweighted by education, age, region, and gender so that the results represent the adult population of Nigeria. Data for this analysis was accessed and constructed on July 21, 2025. The authors had no access to personally-identifying data about survey participants at any point during or after data collection.

In this paper we calculate summary statistics and we use logistic regression to identify factors associated with health insurance coverage, with care utilization, and with use of different levels of care (e.g. public primary care versus private sector or hospital care). We present access to care by insurance categories (NHIS, private insurance, no insurance) graphically, including means by category and 95% confidence intervals. We report full multivariate logistic regression output including control variables in the Supplementary Information where indicated. These regression models include controls for urban/rural residence, gender, education, age categories, region, and income terciles.

Outcome variables include usual source of care (level and ownership of facilities); receipt of population-level screenings in the past 12 months (blood pressure, blood cholesterol, blood sugar, eyes and dental screenings); self-reported health (1-5 scale); whether the respondent had any chronic illness (0/1); and the number of health facility visits and inpatient visits in the past 12 months.

Health insurance status was measured by the questions: “Do you have health insurance? Health insurance is a plan that helps cover the cost of healthcare when you need it,” and “What type of health insurance do you have? If more than one, please tell us the primary type of health insurance you use.” Response options for Nigeria included NHIS, private insurance, company-provided health insurance, community-based health insurance (CBHI), and other. Private and company provided insurance were recoded as a single “private insurance” category for this analysis.

### Ethics statement

This analysis was determined to be not human subjects research by the Harvard T.H. Chan School of Public Health Institutional Review Board (protocol 25-0781), as the research was secondary analysis of a publicly available, fully anonymized dataset.

## Results

The People’s Voice Survey sample is 49% female, with a mean age of 36. 64% of respondents live in urban areas, and 13% have a secondary education or higher.

In this population, access to any form of health insurance is low (7%). The most common form is private insurance (3.6%), followed by public/NHIS (1.7% of the population), with community-based insurance and other forms of insurance reported by less than 2% of the population. (Table 1).

**Table 1:** Sample characteristics.

|  |  |
| --- | --- |
| Observations | N = 2,555 |
| Age | 36.3 (13.9) |
| Female | 1,260 (49.3%) |
| Secondary or higher education | 341 (13.3%) |
| Urban | 1,624 (63.6%) |
| Number of health visits, past year | 2.0 (2.5) |
| Insured | 185 (7.2%) |
| NHIS | 43 (1.7%) |
| Private insurance | 93 (3.6%) |
| Has usual source of care | 1,841 (72.1%) |
| Has chronic condition | 320 (12.5%) |
| Self reported health (1-5) | 2.9 (0.9) |
| Continuous variables presented as: mean (SD) |  |

Factors positively associated with private insurance coverage include post-secondary education, higher income, age group 40 to 49, and residence in south-west zone. Factors positively associated with NHIS coverage are high income, secondary education, age categories 40-49 and 50-59, and residence in north-west zone, while residence in southwest zone is negatively associated with NHIS coverage. (Figure 1). The largest association is between higher education and insurance, with OR 3.6 (95% CI 2.4, 5.3) for private insurance and OR 5.86 (95% CI 3.0, 11.3) for NHIS.

**Figure 1:**
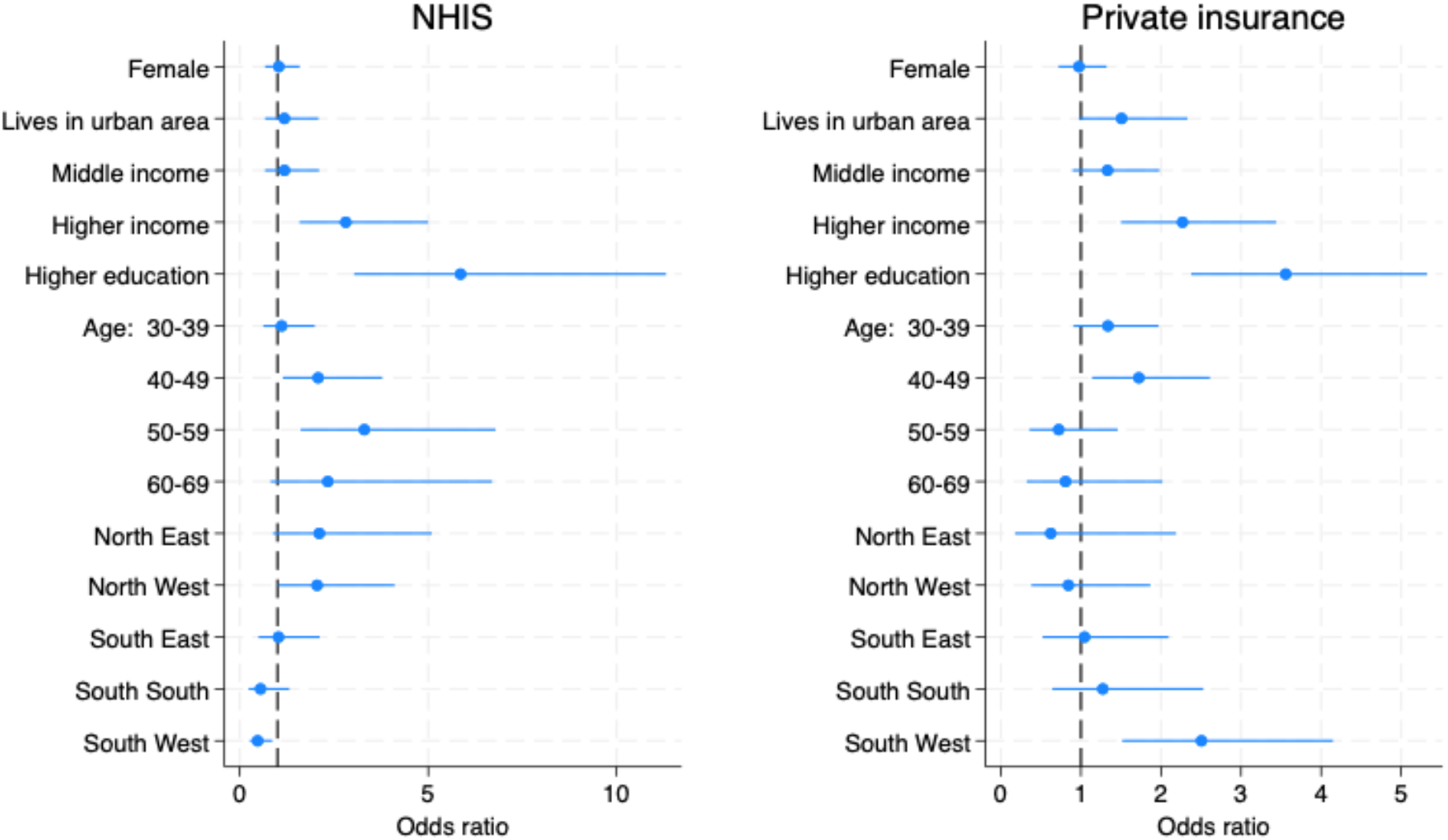
Factors associated with having either NHIS or private health insurance.

Overall, the use of public health care and of primary care as a usual source of care in Nigeria is low. While seventy-two percent of Nigerians report having a usual source of care (Table 1), only 20% of these individuals access care from public primary care providers. In contrast, 38% rely on public secondary providers and 30% use private secondary providers. This pattern is even more pronounced among those who have insurance. Individuals with both public and private insurance are less likely to use public primary providers, and are more likely to use private secondary providers as their usual source of care: 2% of individuals with NHIS and 0% with private insurance use public primary care as their usual source, versus 15% of those with no insurance. By contrast 41% of those with NHIS and 47% of those with private insurance use private secondary care as their usual source of care, compared to 21% of those with no insurance (Figure 2). Similar patterns are present when respondents are asked about their last visit to a health facility, rather than about their usual source of care.

**Figure 2:**
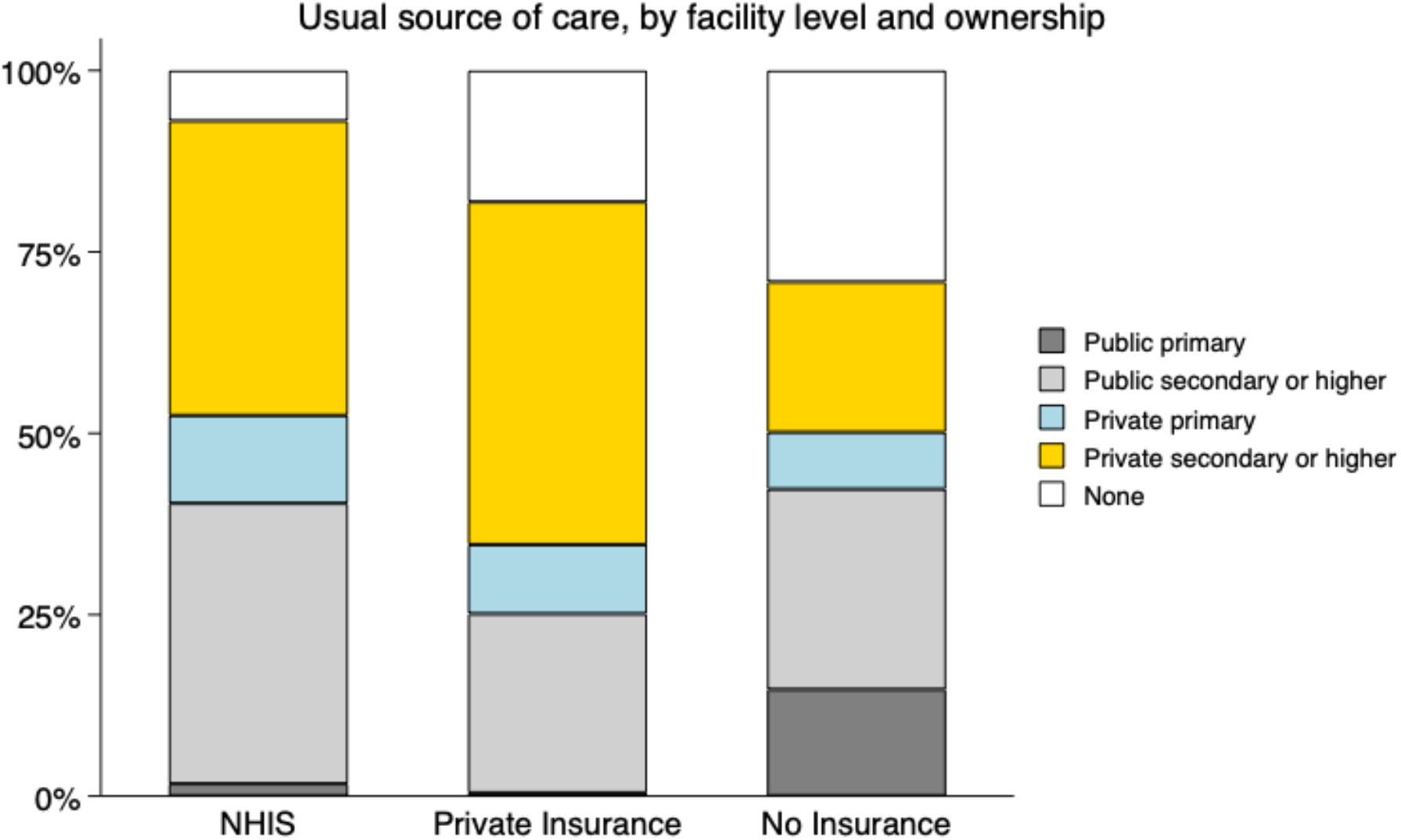
Usual source of care, by facility level and ownership.

In a multivariate regression framework, having any form of insurance is negatively associated with using public primary care (Appendix Table A.1), including when controlling for age, education, gender, urban/rural residence, income tercile, and region of residence (OR 0.48; 95% CI 0.27-0.85).

Respondents with private insurance and NHIS insurance are more likely to have received key screenings than those without insurance: for blood pressure, the figures for respondents over age 40 are NHIS (93%) vs private (72%) vs no insurance (63%). For blood sugar, NHIS had highest screening rates (75%) followed by private (74%), then no insurance (44%). For cholesterol checks, screening rates are as follows: NHIS (26%), private (30%), and no insurance (18%)(Figure 3). Overall, those with NHIS insurance were most likely have had their blood pressure taken, while rates of blood sugar and cholesterol screening are similar between NHIS and private insurance. For all three metabolic screenings, multivariate regressions which adjust for age, income, education, region, gender, and urban/rural residence show that those with any form of insurance had higher screening rates than those without insurance (p<0.01). (Table A2)

**Figure 3:**
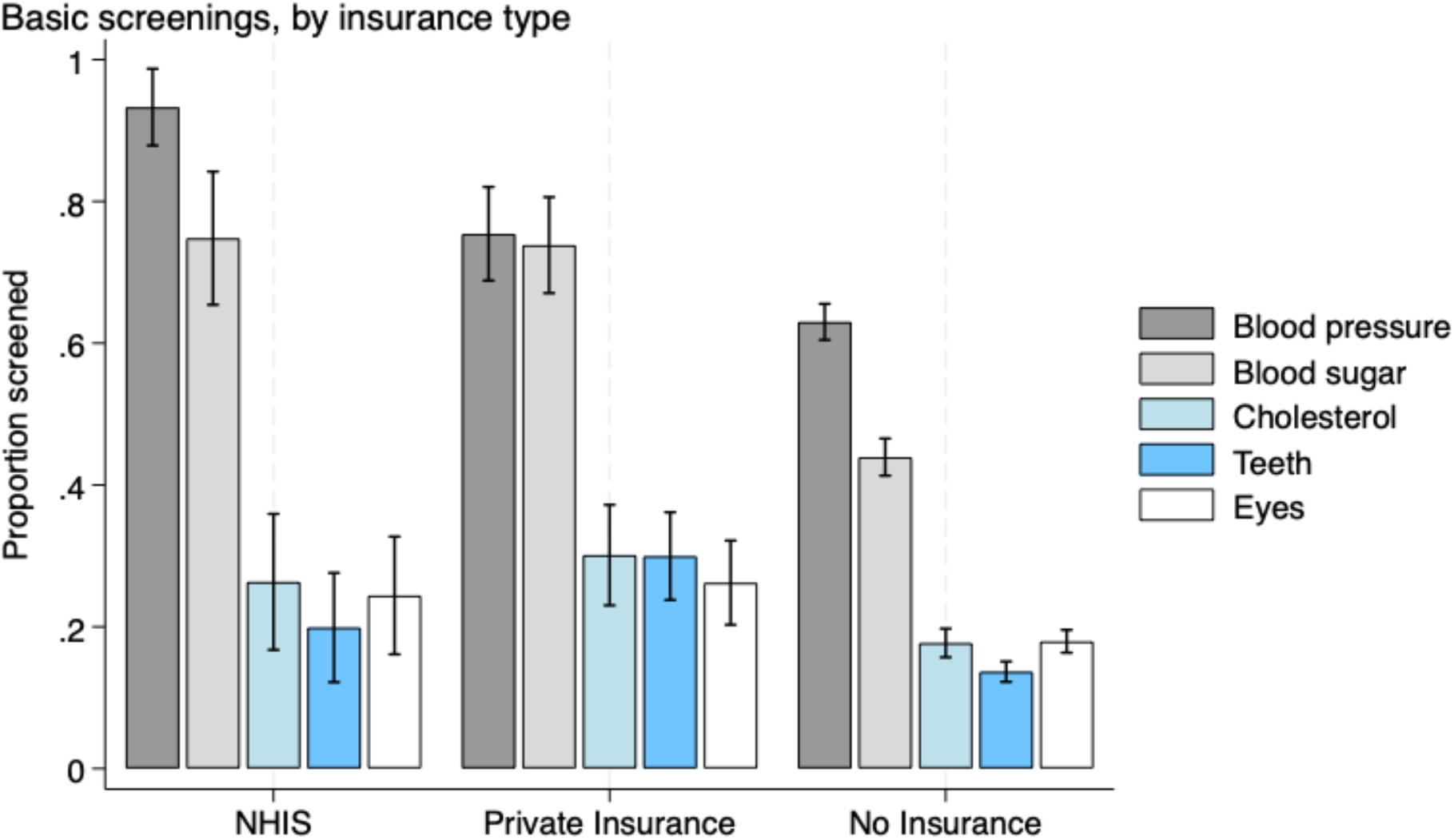
Basic screenings received in the past year (among screening-eligible respondents), by insurance type. Note: for blood pressure, blood sugar, and cholesterol exams, respondents over age 40 are considered screening eligible. For eye and teeth exams, all respondents are considered screening eligible

We next consider eye exams and dental screenings, for the full surveyed population rather than restricting the sample to respondents over 40 years of age. 20% of those with NHIS, 30% of those with private coverage, and 14% of those with no insurance had their teeth checked in the past year. 24% of respondents with public insurance (NHIS), 26% of those with private insurance, versus 18% of those with no insurance had their eyes or vision checked at any point in the last year. For both eye and dental check-ups, multivariate regressions which adjust for age, income, education, region, gender, and urban/rural residence demonstrate that those with any insurance had higher screenings rates than those without insurance (p<0.01). (Table A2)

A frequent challenge for non-mandatory and non-universal health insurance programs is that of adverse selection; i.e. individuals are more likely to purchase health insurance if they know that they are in poor health and likely to need large amounts of medical care in the future. This approach is not expected to be a first order concern in settings with limited voluntary enrollment. Accordingly, in Nigeria it does not seem that individuals with either NHIS or private insurance have worse self-reported health or more chronic conditions than those no insurance (Figure 4). Mean self-reported health (1-5 scale) is 3.9 for those without insurance, 3.7 for those with NHIS, and 4.0 for respondents with private insurance. Prevalence of any chronic condition is 12% (NHIS), 12% (private insurance), and 13% (no insurance). Multivariate regressions with insurance status as a predictor of health status (controlling for age, income, education, region, gender, and urban/rural residence) show that there is no statistically significant relationship between these outcome variables and access to health insurance (Appendix Table A.1).

**Figure 4:**
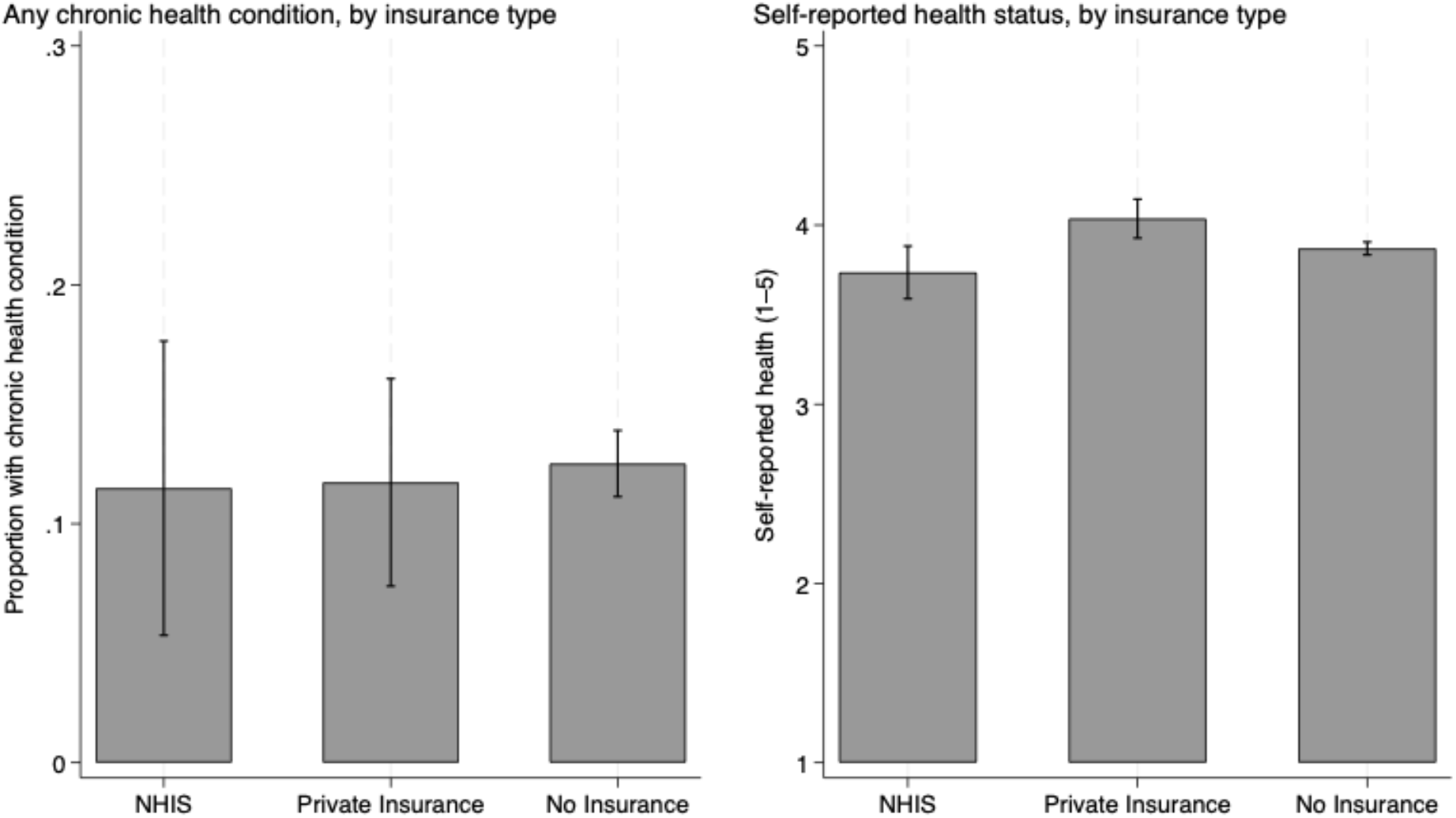
Self reported health, by insurance status.

**Figure 5:**
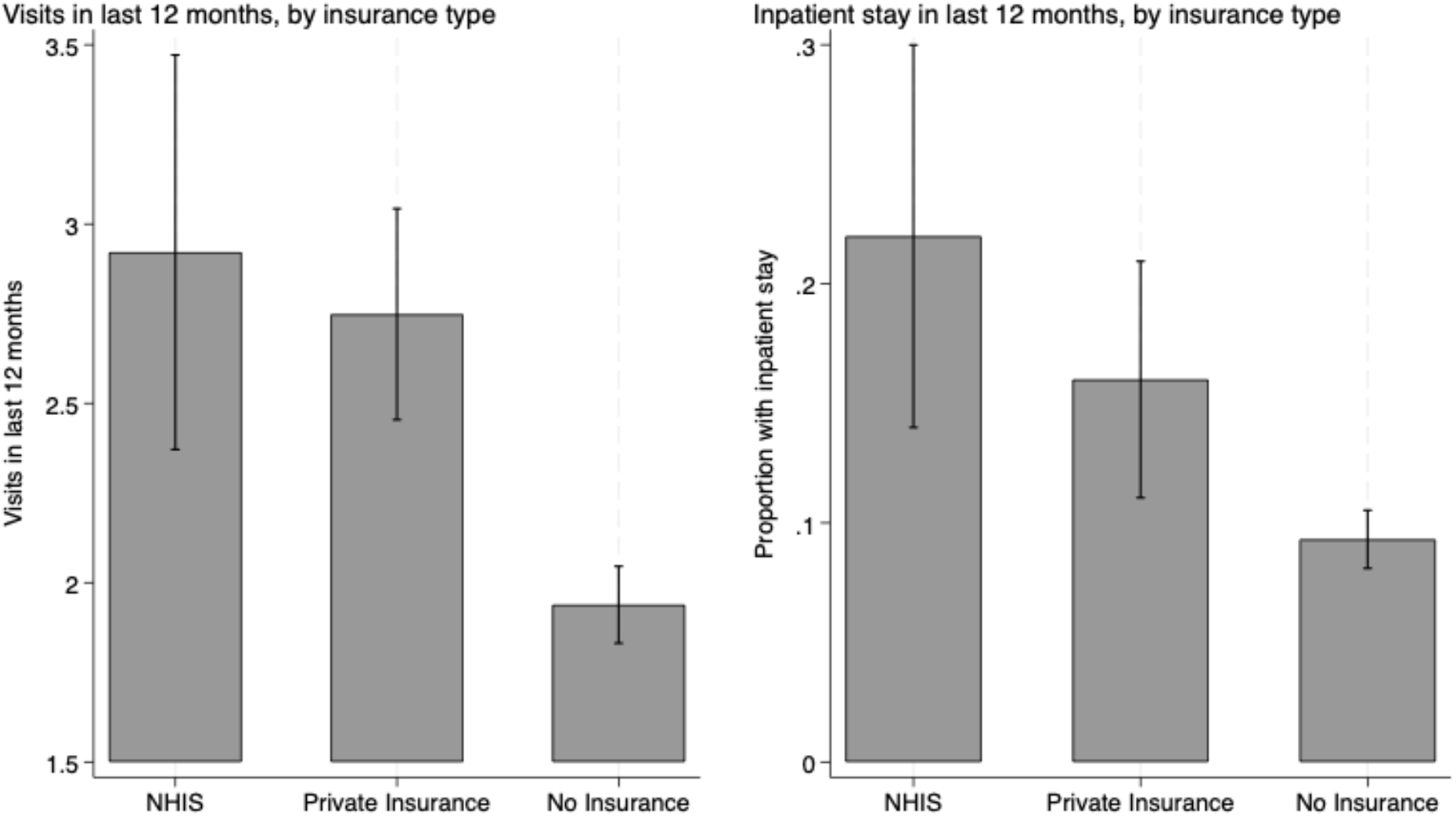
Access to care in last 12 months, by insurance type.

Next we examine care utilization as a function of insurance coverage. (Figure 4). NHIS holders have the highest average number of visits (2.9) and are most likely to have had an inpatient stay in the past 12 months (22%), followed by those with private insurance (2.7 annual visits, 16% inpatient stay). Those with no insurance have the fewest number of visits on average (1.9) and are least likely to have had an inpatient stay (9%). In a regression framework, respondents with any form of insurance access more care and are more likely to have had an inpatient episode than those without insurance (p<0.01) (Table A.3).

## Discussion

In this paper we use a new data source to shed light on an important policy issue in Nigeria and in LMICs more broadly: the population-level coverage of health insurance, and the relationships between coverage and access to care. Despite recent reforms including the 2014 National Health Act and the 2022 National Health Insurance Authority Act, there is still relatively limited research on the scope and distribution of insurance coverage in Nigeria, and the benefits of insurance for enrollees. We can group our main findings into three categories.

Using the PVS, we observe low health insurance coverage overall, and low coverage of the public health insurance scheme (NHIS) at the time that the survey was conducted in mid-2023. Furthermore, these results indicate that public sector insurance coverage is not well-targeted to poorer individuals: higher education and higher income levels are the best predictors of NHIS coverage. This reinforces previous findings on the exclusion of rural and vulnerable populations ^21^. Weak association between existing schemes and utilization of public PHCs further highlights limitations of these models in their current form to rapidly drive uptake of pro-poor, high impact interventions – a central tenet of the UHC agenda.

Second, the effect of insurance on access to care is another focus of our study. While those with insurance do not appear to be in worse health than those without, they do access more care. It is perhaps more notable *where* they access this care. Although robust primary health care systems are widely considered to be critical to enable countries to achieve universal health care and the SDGs, in PVS data we observe low utilization of primary level care as usual source of care, while those with insurance are more likely to access secondary and tertiary care as their usual source. This is likely linked to the finding from previous research that PHCs in Nigeria are under-resourced, poorly maintained, and often do not have the complement of staff required. These issues have implications for how the beneficiaries perceive the quality of care delivered.^22^

Insurance enrollees are also more likely to have received prevention and health promotion services for chronic and non-communicable diseases. These associations may indicate better financial access and better understanding of how to navigate the health system by the insured. The skewed utilization pattern towards private secondary facilities indicates that insurance, in its current form, may be reinforcing rather than correcting the imbalances and inefficiencies of the system.

The data does not support concerns of adverse selection into insurance based on poor health status or chronic illness. This is likely due to low voluntary enrolment rates, higher enrolment of often healthy individuals in the organized private sector, and/or administrative barriers to enrolment even among those with high need, underscoring the importance of mandatory coverage provisions now embedded in the NHIA Act. Thus, the fact that enrollees are not less healthy than the uninsured may not be surprising. It does however highlight the inequities reflected in their use of both more preventive and curative health services than the insured.

Our findings are largely consistent with previous literature. For example PVS results from Ethiopia, Kenya, South Africa, India, and Laos showed lower health utilization and access to preventive screenings among the uninsured population relative to those with private insurance, and reduced utilization and access for those with public relative to private insurance.^19^ Our estimates of low population coverage of health are also consistent with previous national population surveys in Nigeria.^23^ Nigeria’s health insurance coverage (7%) is broadly consistent with trends across sub-Saharan Africa where access to health insurance has been estimated at 17% on average.^24^

We also highlight several relationships to ongoing policy initiatives, as well as implications for policymakers. Many African countries have scaled up insurance programs in an effort to achieve UHC goals associated with the SDGs, and ultimately improve access and affordability. However, in many settings, evidence suggests that current public insurance programs are insufficient.

Community based health insurance and social health insurance generally have been found to have positive effects on service utilization,^25^ which also which aligns with our findings. For example, Rwanda, South Africa, and Ghana have seen positive effects of national health insurance on service use.^26–29^ At the same time, countries in sub-Saharan Africa that have implemented national health insurance programs have not clearly outperformed their neighbors,^30^ there is limited evidence that they have improved financial protection or increased health expenditure,^31^ and effects on quality of care received, and ultimately on population health, have not been conclusively documented in existing literature.^5^

We also note several distinct strengths and limitations of paper. The PVS captures individuals’ perspectives and experiences which are often overlooked in health care utilization data, analysis, and thus policy decisions. The opinions of health system users and citizens are key to system legitimacy and trust, and it will likely be valuable to incorporate these human elements with other forms of service delivery data to inform policy decisions. However, we acknowledge potential sampling bias of phone-based surveys; despite reweighting strategies designed to produce population-representative estimates, imprecisions in the reweighting process could result in underrepresentation of groups with limited phone access. In addition, we note that as we are using an observational research design with cross-sectional data, we cannot interpret any reported associations as causal.

The design of national health insurance systems, the quality of implementation, and the political will to support and finance programs over time are critical for NHI to contribute to national health system progress. Nigeria’s recent health reforms offer significant promise, and there are indicators of progress since the time of this data collection in 2023: According to NHIA as of December 2024, there are now 19.2 million Nigerians enrolled on the NHIS.^32,33^ Yet just increasing coverage will not be enough: next steps will require political will and consistent implementation, and ongoing monitoring and adjustment as lessons are learned. Improvements in financing, strategic purchasing, and quality of care will need to accompany increased demand for, and utilization of, services.

## Data sharing statement

People’s Voice Survey data is publicly available upon registration from https://dataverse.harvard.edu/dataverse/pvs.

## Acknowledgements

We would like to acknowledge Prof. Todd Lewis for assistance with access to the People’s Voice Survey data.

## Supporting information table legends

**Table A1: Association between insurance, health utilization, and status (Logistic regression)**

**Table A2: Association between insurance and preventive screenings (Logistic regression)**

**Table A3: Association between Insurance and Facility Visits (Logistic regression)**

